# Model stability of COVID-19 mortality prediction with biomarkers

**DOI:** 10.1101/2020.07.29.20161323

**Authors:** Chenyan Huang, Xi Long, Zhuozhao Zhan, Edwin van den Heuvel

## Abstract

Coronavirus disease 2019 (COVID-19) is an unprecedented and fast evolving pandemic, which has caused a large number of critically ill patients and deaths globally. It is an acute public health crisis leading to overloaded critical care capacity. Timely prediction of the clinical outcome (death/survival) of hospital-admitted COVID-19 patients can provide early warnings to clinicians, allowing improved allocation of medical resources. In a recently published paper, an interpretable machine learning model was presented to predict the mortality of COVID-19 patients with blood biomarkers, where the model was trained and tested on relatively small data sets. However, the model or performance stability was not explored and assessed. By re-analyzing the data, we reveal that the reported mortality prediction performance was likely over-optimistic and its uncertainty was underestimated or overlooked, with a large variability in predicting deaths.

---

We read with great interest the recently published article by Yan et al.^1^ on an interpretable and highly needed mortality prediction model for COVID-19 patients. We commend the authors’ initiative to timely look into mortality for this unprecedented and evolving global health crisis. Based upon a database of blood test samples from 485 infected patients (with multiple blood samples per patient) in the region of Wuhan, China, a tree-based machine learning model was employed to predict the outcome of individual patients (death/survival). Three biomarkers (features) were experimentally selected, including lactic dehydrogenase (LDH), high-sensitivity C-reactive protein (hs-CRP) and lymphocyte, with good predictive values of disease deterioration or fatality proven in previous studies^2-5^. The authors claim that the model can predict the outcome approximately 10 days in advance with an accuracy of >90%.

According to the paper and source code by Yan et al.^1^, the three important features were selected based on a training data set of 375 patients using a 100-round five-fold cross-validation and XGBoost decision-trees. To empower early identification of COVID-19 mortality, they considered only blood samples with complete measurements of the three features. Therefore, with the three selected features, a ‘single-tree XGBoost’ model was re-trained on a single subset (70%) of 351 patients and validated on the remaining 30% of the training data. The resulting model was then tested on an external dataset with 110 patients.

We believe that both the training (including validation) and test data sets are relatively small, indicating a potential concern with regards to the precision and stability of the reported prediction model, especially when the test dataset contains only 13 deaths out of 110 patients. We are concerned that the published results are an over-optimistic estimation of true prediction performance^6^, in particular when training the model is conducted with a single run without cross-validation. Therefore, this letter aims at addressing the matter of potential model instability in predicting mortality for COVID-19 patients, by demonstrating high variability of prediction results.

We run 1000 times the model training and validation (7:3 random split) using the same three features and the same single-tree method as Yan et al.^1^, and evaluated the variability of prediction results on the external test dataset with 110 patients. We applied our approach to both the 110 latest complete samples used by Yan et al.^1^ as well as the 251 complete blood samples. Boxplots^7^ of the four performance metrics used by the authors over our 1000 runs are presented in Fig. 1.

**Fig 1.**
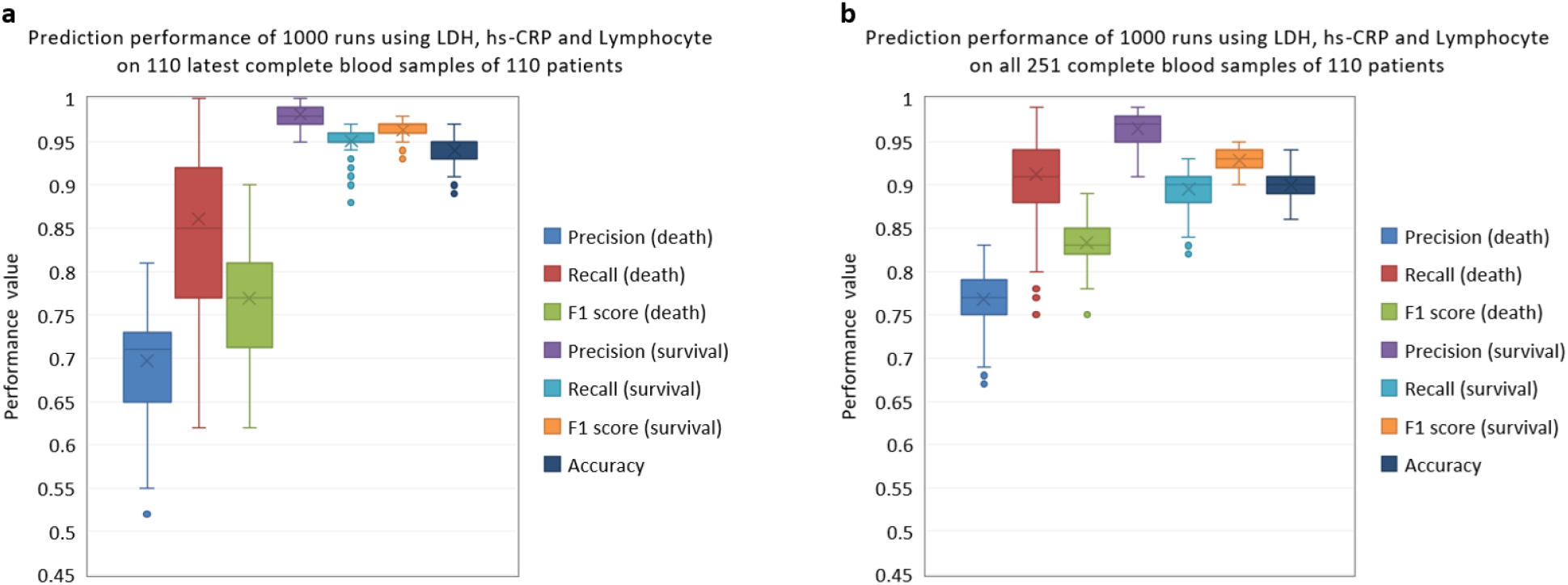
Prediction performance of 1000 runs on data from 110 patients in the external test set using the three selected features (LDH, hs-CRP and lymphocyte). **a**, Boxplots of prediction results on 110 latest complete blood samples (13 samples corresponding to deaths). **b**, Boxplots of prediction results on all 251 complete blood samples (69 samples corresponding to deaths). Note that, for each boxplot, the line within the box, the cross mark, the box and the error bars indicate mean, median, quartiles and whiskers, respectively, and the dots correspond to outliers.

As shown in Fig. 1a, the precision, recall and F1 score^8^ have a small variability in predicting survivals, relatively consistent with the original results^1^. However, they all show a large variability in predicting deaths (mean precision of 0.70 ranged from 0.52 to 0.81, mean recall of 0.86 from 0.62 to 1, and mean F1 score of 0.77 from 0.62 to 0.9) than reported in the original paper^1^, with an obvious discrepancy between training and test results. Similar findings can be observed in Fig. 1b when testing on all the 251 blood samples, despite a slightly better performance in predicting deaths (likely due to a more balanced distribution in terms of death and survival numbers), but at the expense of a reduced prediction in survivals.

For the analysis of prediction horizon, predicting the clinical outcome in a timely manner can provide early warnings to clinicians, allowing improved allocation of medical resources. Yan et al.^1^ applied the trained single-tree model on the 251 blood samples and analyzed the model performance over time for different days to outcome. To visualize the variability on days of prediction upfront, Fig. 2 illustrates the death prediction performance over the runs versus days to outcome (from 0 to 23 days). A remarkable difference across the 1000 models can be noticed in different “days to outcome”, even when close to the day of outcome (day 0). For example, the recall at 1 day in advance of outcome ranged from 0.67 to 1, indicating that there was a chance of training a prediction model that can miss, on average, 14%, and up to 33% of the actually death samples, unlike the result published by Yan et al.^1^ with an F1 score of 1. This suggests an evident overly optimistic view of results in identifying mortality for and over different days in advance and disputes the claim of accurately detecting deaths already around 10 days before death.

**Fig 2.**
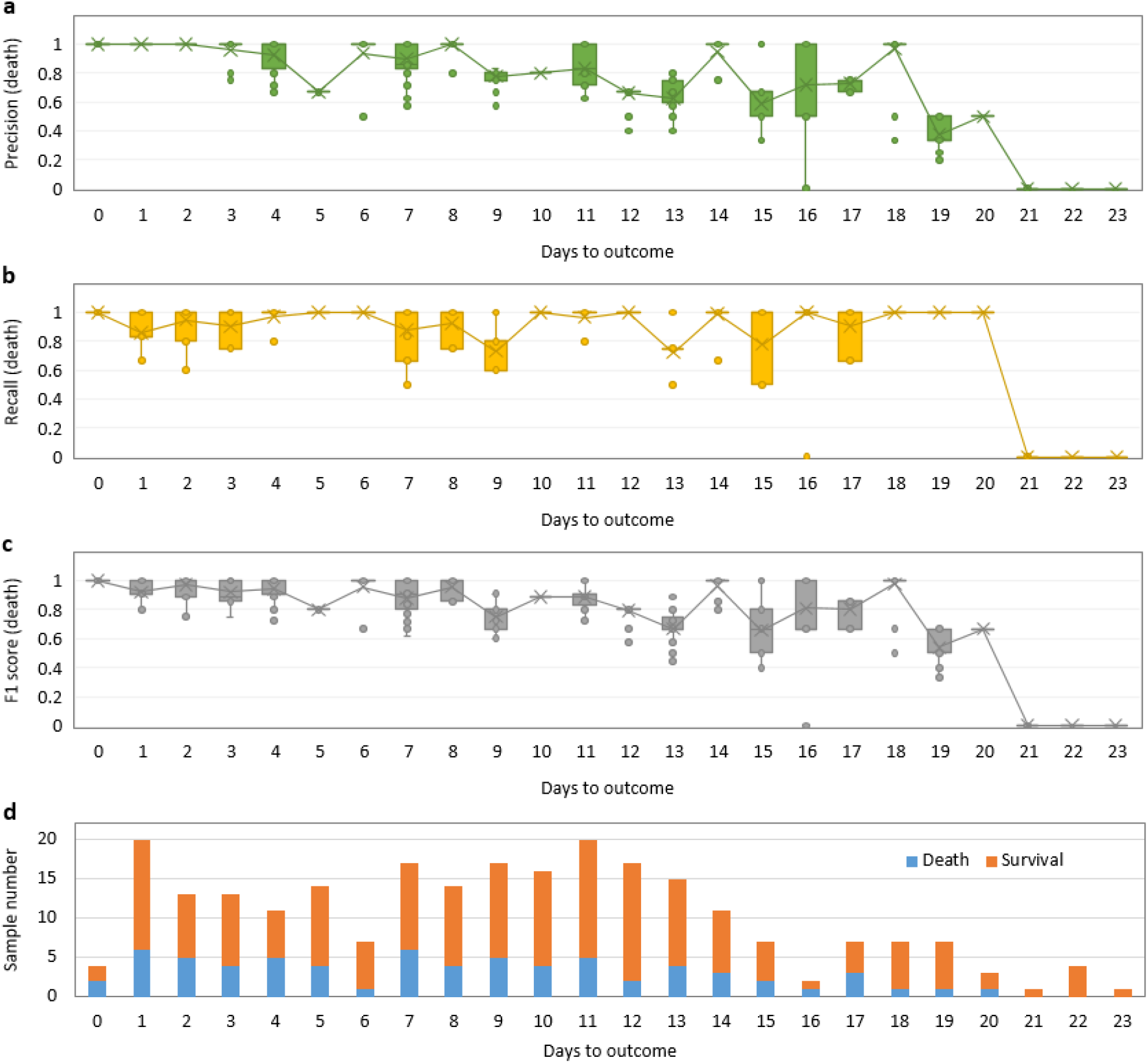
Prediction performance of 1000 runs with respect to the days to outcome for 110 patients (251 complete blood samples) in the external test set. **a**, precision, **b**, recall and **c**, F1 score in death prediction, and **d**, the corresponding sample numbers of death and survival samples.

The root causes contributing to the demonstrated instability or uncertainty of the prediction results could be twofold. First, the modelling could be overfitting to the training data that is of a small size (without cross-validation) and likely not representative of the entire patient population in terms of the characteristics of the biomarkers from disease onset to death as well as death cause^9,10^, not generalizable to external unseen data. Second, the external test dataset is too small (in particular on different days to outcome) to draw a firm conclusion regarding model stability. In addition, we found a discrepancy in class imbalance between the training (159 death cases out of 351 patients) and the test dataset (13 death cases out of 110 patients). The percentage of deaths in the training set was far from the actually fatality rate of COVID-19 (varying from 1.4%^11^ to 4.3%^12^ in different regions and hospitals, and higher in critical patients). As a result, the trained model would be more likely to predict as death cases compared with the intrinsic prior probability of death (i.e. fatality rate), resulting in a higher recall while a lower precision^13^, in particular when using ensemble machine learning models not designed for data with a strongly skewed class distribution^14^ including XGBoost^15^. Therefore, with the existent data used in the study, model stability should be carefully considered.

Although the study by Yan et al.^1^ shows a recognizable promise in predicting mortality for COVID-19 patients with three biomarkers using a single-tree XGBoost model, we reveal that the reported prediction performance was over-optimistic. The prediction results remain unstable (exhibiting large variabilities) and therefore the assertions made by Yan et al.^1^ on the effective days to predict in advance and the corresponding accuracy seem not sufficiently solid, not fully supported by the evidence presented in this article. As the authors discussed as well, a larger representative cohort of COVID-19 patients is imperatively required to further verify the performance and stability of the proposed mortality prediction model in the future.

## Data Availability

Publicly available data. The data were retrieved from the supplementary information of the published work by Yan et al. (https://doi.org/10.1038/s42256-020-0180-7) on May 16, 2020.

https://static-content.springer.com/esm/art%3A10.1038%2Fs42256-020-0180-7/MediaObjects/42256_2020_180_MOESM3_ESM.zip

## Data and code availability

The data and code used herein were retrieved from the supplementary information of the published work by Yan et al.^1^ on May 16, 2020.

## Author contributions

C.H. and X.L. performed the analysis. X.L., C.H., and Z.Z. contributed to the text. X.L. and E.vd.H. interpreted the results. All authors provided critical review and approved the manuscript.

